# Smartphone addiction and its association with common mental disorders among students attending the university of Dschang, West region, Cameroon

**DOI:** 10.1101/2020.02.29.20029405

**Authors:** Martial Pianta Sonkoue, Benjamin Momo Kadia, Miranda Baame Esong, Cyrielle Djouda Douanla, Jerome Ateudjieu

## Abstract

**Background:** Smartphones are an integral part of modern communication but addiction to these devices could predispose to common mental disorders.

**Objectives:** To determine the prevalence of smartphone addiction and common mental disorders as well as the association between smartphone addiction and these mental disorders in Dschang university students who use smartphones.

**Methods:** A cross-sectional study was conducted. Randomly selected students completed a survey comprising Patient Health Questionnaires seven and nine (PHQ-9 and PHQ-7) to screen for depression and anxiety respectively, and the Smartphone Addiction Scale-Short Version (SAS-SV) to grade smartphone use. The association between smartphone use and common mental disorders was assessed using logistic regression modelling.

**Results:** We recruited 634 participants. The prevalence of smartphone addiction was 20.98% [C.I:17.99%-24.32%]. The prevalence of depression and anxiety were 34.86% [95% CI: 31.25-38.65] and 19.27% [95% CI: 16.81-22.99] respectively. The odds of depression in students with addiction was 5 times the odds of depression in students without addiction [OR: 4.96, 95% CI: 3.30-7.45, p<0.001]. The odds of anxiety in students with addiction was 2 times the odds of anxiety in students without addiction [OR: 2.44, 95% CI: 1.58-3.77, p<0.001]. After adjusting for age, sex, faculty, cycle of study, marital status, religion, chronic diseases, sexual abuse and suicidal ideation, there remained a statistically-significant association between smartphone addiction and both depression [OR: 4.46, 95%CI; 2.92-6.84, p<0.001] and anxiety [OR: 2.08, 95% CI; 1.31-3.30, p=0.002].

**Conclusion:** In this setting, it is crucial to foster strategies of prevention and promotion in mental health especially among problematic smartphone users.

## 1. Background

Common mental disorders refer to two main diagnostic categories: depressive disorders and anxiety disorders [1]. These disorders are highly prevalent in the general population (reason why they are considered ‘common’), and impact on the mood or feelings of affected persons [1]. Symptoms range from mild to severe in terms of severity and months to years in terms of duration [1]. These disorders lead to huge losses in work participation and productivity, and yet lend themselves to effective and accessible treatment as part of an integrated program of chronic disease management [1]. In 2010, worldwide, an estimated US$2.5–8.5 trillion in lost output was attributed to mental, neurological and substance use disorders, depending on the method of assessment used [2]. This amount is expected to nearly double by 2030 if a concerted response is not mounted [2]. In view of this concern, the promotion of mental health and wellbeing has been explicitly included in the United Nations’ 2015–30 Sustainable Development Goals [3]. A review reported a prevalence of depression of 33% among university students [4] while two other reviews on anxiety reported a global prevalence ranging from 0.9-28.3% [5] and a prevalence of anxiety disorders in adult populations of 3.8-25% [6] respectively. Student mental health is a growing concern [7] and there is evidence that university students tend to have higher rates of depression than the general population [4].

A smartphone is a mobile phone that performs many of the functions of a computer, typically having a touch screen interface, internet access, and an operating system capable of running downloaded applications [8]. It had been projected that by 2020 the population of smartphone owners would reach 6.1 billion, which is about 70% of the world’s population [9]. Many new users are expected to emerge from developing countries through greater device affordability, growing economies, and young, growing populations [9]. Several studies have examined the relation between smartphone use and common mental disorders [10,11,12,13,14]. Without statistically adjusting for other relevant variables, depression severity was consistently related to problematic smartphone use [15]. The extent of the problem of depression and anxiety among students in low-and middle-income countries (LMICs) is largely unknown [16]. In a recent systematic review with meta-analysis, Sohn and colleagues examined the association between problematic smartphone use and mental health outcomes amongst children and young adults. Although this was a global review which included evidence for the seven-year period 2011-2017 and had no language restriction in its literature search strategy, only studies from Europe, America and Asia were retained. This strongly indicates the huge gap in evidence on the association between smartphone addiction and common mental illnesses in sub-Saharan Africa [17]. This study therefore sought to estimate the prevalence of smartphone addiction in university students who use smartphones; to estimate the prevalence of common mental in the students and to determine the association between smartphone addiction and these mental disorders.

## 2. Methods

### 2.1. Ethical considerations

Ethical registry was granted by the Cameroon National Committee for Research Ethics for Human Health. The registry number is 2018/07/1081/CE/CNERSH/SP.

### 2.2. Study design and period

This was an analytic cross-sectional study conducted from the 10 March to the 30 April 2018.

### 2.3. Study area and setting

The study was conducted on the main campus of the university of Dschang. The university of Dschang is one of the eight public universities of Cameroon. It is a bilingual university located in the West region of Cameroon in Menoua division within the town of Dschang and had a population of about 32000 students in 2018. The university of Dschang has six faculties located on the main campus (Faculty of Science, Faculty of letters and human sciences, Faculty of economics and management, Faculty of law and political sciences, Faculty of agronomy and agricultural sciences, faculty of medicine and pharmaceutical sciences) and two institutions located out of the main campus (Foumban Institute of fine arts and Fotso Victor institute of Bandjoun). The faculty of medicine and pharmaceutical sciences was newly created by a presidential decree and went functional during academic year 2017 /2018. A small guidance counseling unit is located at the rectorate. Even though it serves the whole university, the unit is underutilized and seldom functional.

### 2.4. Study participants and selection criteria

Students from the University of Dschang who possess and use smartphones during the 2017 and 2018 academic year. The following selection were set:

#### Eligibility criteria

Being a registered student of the university of Dschang in the academic year 2017/2018 who uses a smartphone.

#### Inclusion criteria

Being a registered student of the university of Dschang in the academic year 2017/2018 who uses a smartphone and consent to participate in the study

#### Exclusion criteria

students with known history of psychiatric disorders; not responding to up to 80% of questions on the questionnaire.

### 2.5. Sampling procedure

A multistage random sampling method was used. Two faculties were selected at random from the six faculties of the University of Dschang (FESM: Faculty of Economic Sciences and Management, FAAS: Faculty of Agronomy and Agricultural Sciences, FS: Faculty of Science, FLPS: Faculty of Law and Political Sciences FLHS: Faculty of Letters and Human Sciences, FMPS: Faculty of medicine and pharmaceutical sciences) and participants were recruited from strata designated by cycle of study in each selected faculty. The strata were the bachelors, masters and PhD study cycles. A systematic review found a prevalence of depression of 33% among students [4]. Using the Cochran’s sample size determination formula

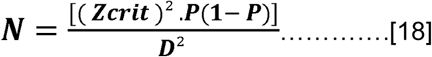

and substituting p=33%, relative precision (D) =0.05, normal standard deviate Zcrit=1.96 at significance criterion of 0.05, we obtained a minimum sample of N=339 students. Adjusting for a non-response rate of 10% and a design effect of 2 we got a final sample size of 758 students.

### 2.6. Variables and data collection

A semi-structured questionnaire comprising PHQ-9, PHQ-7 and SAS-SV was conceived by the principal investigator and pre-tested on 50 students from the Faculty of Medicine and Biomedical Sciences and the Faculty of Science of the University of Yaounde 1. Participants were students from the Faculty of Agronomy and Agricultural Sciences (FAAS) and the Faculty of Economic Sciences and Management (FESM). After explaining the purpose of the study, participants’ consent was obtained before they responded to the self-administered questionnaire consisting of sociodemographic characteristics, PHQ-9 and PHQ-7, Smartphone use and SAS-SV. A case of depression was defined as a Patient Health Questionnaire-9 (PHQ-9) score of 11 or more. A case of anxiety was defined as a Patient Health Questionnaire-7 (PHQ-7) score of 11 or more. A cut off point of 11 was used for PHQ-9 and PHQ-7 to screen for depression and anxiety respectively. A cut-off score for the PHQ-9 of 11 had the best trade-off between sensitivity (89%) and specificity (89%) [19].

Smartphone use was the exposure of interest. It was assessed using the Smartphone addiction scale short version (SAS-SV) to distinguish excessive smartphone users (smartphone addicted) from normal smartphone users (not addicted). The smartphone addiction scale is a self-diagnostic scale used to identify ‘smartphone addicts’ [20]. This tool has been reduced to create a short version (SAS-SV) [20] which was eventually adapted to French [21]. The SAS-SV is a validated scale originally constructed in South Korea but published in English. It contains ten items rated on a dimensional scale (1= ‘strongly agree’ to 6= ‘strongly disagree’). The total score ranges from 10 to 60 with the highest score being the maximum presence of ‘smartphone addiction’ in the past year. Kwon and colleagues proposed cut off points per gender: in males it is 31 with a sensitivity value of 0.867 and specificity value of 0.893 and in females it is 33 with sensitivity value of 0.875, and a specificity value of 0.886 [20]. In this study, we used the mean i.e, 32 out of 60 as cut off for both genders to identify ‘excessive smartphone users’ (smartphone addicts) since no statistical difference was found between SAS-SV total score per gender [21].

### 2.7. Statistical analysis

Data were analysed using Epi-info version 7.2.2.6 statistical software. Descriptive statistics (mean, frequencies and percentages) were used to report participants demographics. PHQ-9, PHQ-7 and SAS-SV scores were categorized using their cut off values and analysed as binary variables. The prevalence of a common mental disorder was estimated by dividing the number of students who owned and used smart phones and had the disorder by the total number of students who owned and used smartphones. We estimated the 95% confidence limits for each prevalence. A logistic regression model was built using a priori confounders namely, age, sex, faculty, cycle of study, marital status, religion, chronic diseases, sexual abuse and suicidal thoughts. The model was used to assess the association between smartphone use and depression and anxiety using a significance criterion of 0.05 level and 95% confidence limits.

## 3. Results

### 3.1. Sample coverage

A total of 758 students were approached for this study. After excluding unanswered and partially answered questionnaires, 634 were enrolled (response rate of 84%) of which 307 were females and 327 were males. Overall, females had a better response rate than males. Males in the bachelor’s degree of FESM and females in PhD cycle had the lowest response rates; 50% and 43% respectively. Table 1 shows the response rates by sex and cycle in each faculty.

**Table 1:** Response rate by sex, faculty, and cycle of study.

| Cycle of study | FESM |  | FAAS |  | Total: X% (a/b) |
| --- | --- | --- | --- | --- | --- |
|  | Male: X% (a/b) | Female: X%, (a/b) | Male: X% (a/b) | Female: X% (a/b) |  |
| Bachelor's | 50% (69/138) | 92% (142/155) | 106% (111/104) | 115% (80/69) | 86% (402/466) |
| Masters | 123% (53/43) | 67% (22/33) | 59% (74/126) | 95% (55/58) | 78% (204/260) |
| PhD | 150% (10/6) | 125% (5/4) | 79% (10/14) | 43% (3/7) | 90% (28/31) |
| Total | 71% (132/187) | 88% (169/192) | 80% (195/244) | 102% (138/134) | 84% (634/757) |

X%=Response rate, a=Number included, b=Number expected, FESM: Faculty of Economic Sciences and Management, FAAS: Faculty of Agronomy and Agricultural Sciences

### 3.2. Characteristics of study participants

The study participants were predominantly males (51.58%) and the age range was 16 to 38 years. The mean age was 22.5 ± 3 years. The majority of participants lived alone in a room (51%). Catholicism was the most practiced religion (58.2%). A minority of participants (9%) had a chronic disease, 0.04% had been sexually abused and 9% had experienced suicidal thoughts within the previous 2 years. Table 2 below shows the characteristics of the study participants.

**Table 2:** Characteristics of study participants.

| Variable | Total (634)<br>n (%) | Female (307)<br>n (%) | Male (327)<br>n (%) |
| --- | --- | --- | --- |
| **Lodging** |  |  |  |
| Cohabiting in a room | 75 (12%) | 40 (13%) | 35 (10.7%) |
| Single in a room | 324 (51%) | 200 (61.2%) | 92(28.1%) |
| Family house | 235 (37%) | 92 (28.13%) | 200(61.2%) |
| **Marital status** |  |  |  |
| Married | 10 (1.6%) | 8 2. (6%) | 2 (0.6%) |
| Divorced | 3 (0.5%) | 3 (1%) | 0 |
| Single | 616 (97.2%) | 292 (95.1%) | 324 (99%) |
| Widow(er) | 5 (08%) | 4 (3%) | 1 (0.3%) |
| **Religion** |  |  |  |
| Animist | 2 (0.3%) | 1 (0.3%) | 1 (0.3%) |
| Pentecostal | 38 (6%) | 17 (5.5) | 21 (6.4%) |
| Adventist | 10 (1.6%) | 2 (0.7%) | 8 (2.5%) |
| Catholic | 369 (58.2%) | 196 (63.8%) | 196 (63.8%) |
| Islamist | 39 (6.15%) | 18 (5.7%) | 21 (6.4%) |
| Jehovah Witness | 7 (1.1%) | 5 (1.6%) | 2 (0.6%) |
| Protestant | 145 (22.9%) | 62 (20.2%) | 83 (25.6%) |
| Other | 24 (3.8%) | 6 (2%) | 18 (5.5%) |
| **Parents' education** |  |  |  |
| Primary | 75 (11.8%) | 33 (10.8%) | 42 (12.8%) |
| Secondary | 285 (45%) | 144 (46.9%) | 141 (43.1%) |
| Higher | 274 (43.2%) | 130 (42.4%) | 144 (44%) |
| **Chronic disease** | 54 (9%) | 17 (6%) | 37 (11%) |
| **History of sexual abuse** | 25 (0.04%) | 20 (6.5%) | 5 (1.5%) |
| **Suicidal ideation**<br>**within previous 2 years** | 57 (9%) | 31 (10.1%) | 26 (8%) |

### 3.3. Prevalence of depression and anxiety

The prevalence of depression and anxiety was 34.86% [C.I: 31.25-38.65] and 19.27% [C.I: 16.81-22.99] respectively. Table 3 shows the distribution of depression and anxiety by sex, age and cycle of study.

**Table 3:** Prevalence of depression and anxiety stratified by sex, age and level of study.

| Variable | Depression |  | Anxiety |  |
| --- | --- | --- | --- | --- |
|  | Prevalence | 95%CI | Prevalence | 95%CI |
| **Sex** |  |  |  |  |
| Male | 28.44% | 23.82-33.56 | 17.13% | 13.43-21.58 |
| Female | 41.69% | 36.31- 47.28 | 22.48% | 18.16-27.47 |

| Level of study |  |  |  |  |
| --- | --- | --- | --- | --- |
| Bachelors | 38.06% | 33.45-42.90 | 20.4% | 16.75-24.61 |
| Masters | 29.41% | 23.26-36.18 | 18.63% | 13.53-24.66 |
| PhD | 28.57% | 13.22-48.67 | 17.83% | 6.06-36.89 |
| Adolescent | 28.18% | 20.02-37.56 | 20.91% | 13.74-29.70 |
| Young adult | 38.86% | 34.02-43.93 | 19.84% | 16.08-24.21 |
| Adult | 30.13% | 23.05-37.98 | 18.59% | 12.82-25.59 |

### 3.4. Smartphone use

Participants reported spending on average 7.2 hours per day on their smartphones (with females spending more time (8hours) than males (6.5hours) and spent an average amount of $1.5 per week on communication credit or data bundles for their phones. Smartphones were mostly used for social media activities (39.3%) and short message services (35.8%). Figure 1 shows the main activities performed when using smartphones.

**Figure 1:**
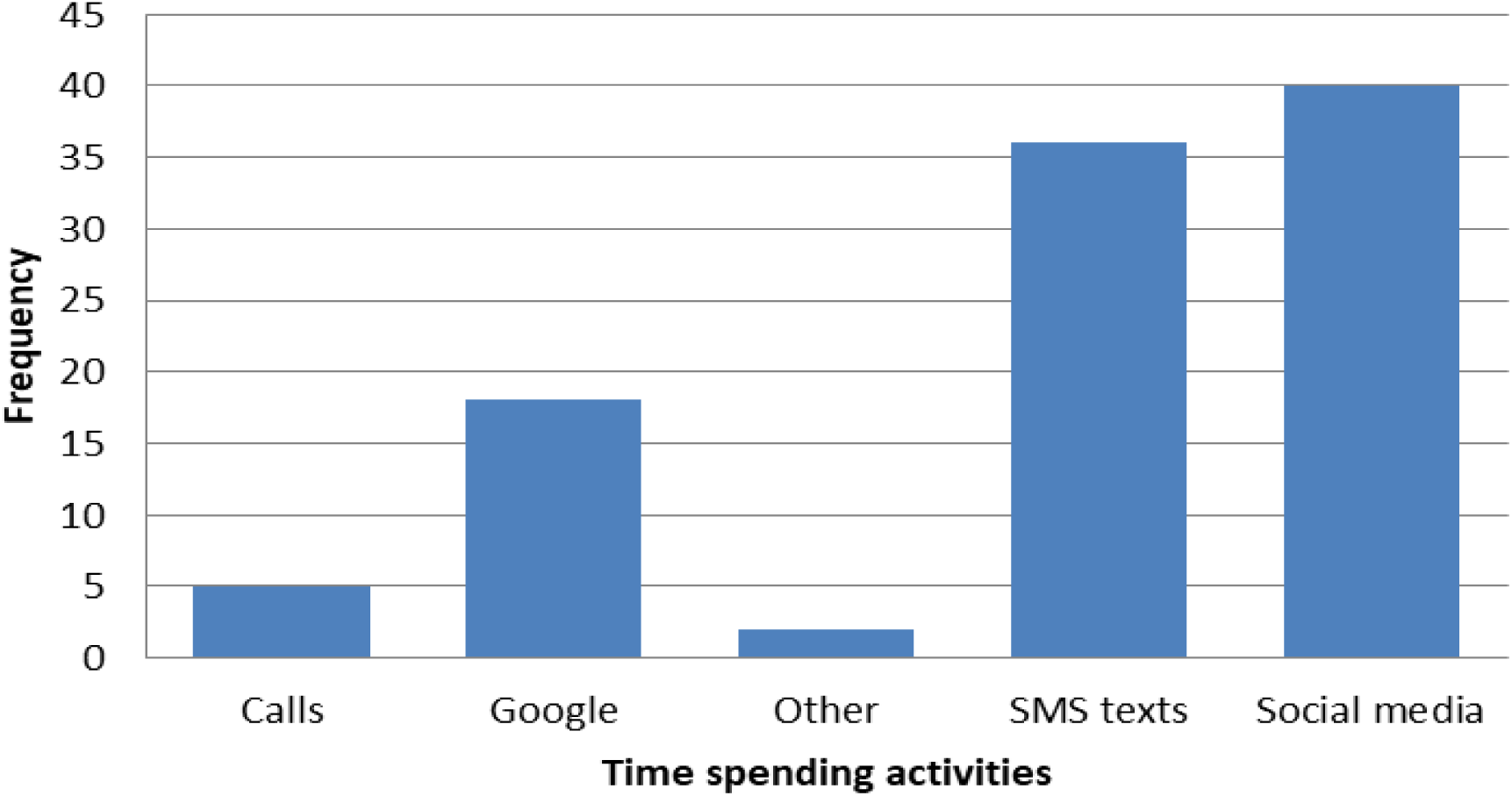
Main activities performed on smartphones by students.

In our sample, 53.67% [340/634, 95% C.I: 49.74%-57.48%] of participants attested that using smartphone affects their academic performance and among these participants, 64.41% [219/634, C.I: 59.19%-69.31%] declared that it affects their performance positively. The prevalence of smartphone addiction was 20.98% [C.I:17.99%-24.32%].

### 3.5. Association between smartphone addiction and depression

The odds of being depressed when addicted to smartphones was 4.96 [95% C.I: 3.30-7.45] the odds of being depressed when not addicted (p<0.001). As shown on table 4, after controlling for age, cycle of study, faculty, matrimonial status, religion, sex, sexual abuse and suicidal thoughts, there was still evidence of a strong association between smartphone addiction and depression (Adjusted Odds Ratio: 4.46, 95% C.I: 2.92-6.84), p<0.001

**Table 4:** Association between smartphone addiction and depression, adjusted for age, sex, faculty, cycle of study, matrimonial status, religion, chronic diseases, sexual abuse and suicidal ideation.

| Variable | Adjusted odds ratio (CI) | P-Value |
| --- | --- | --- |
| **Addiction** |  |  |
| Addicted | 4.46(2.92-6.84) | <0.001 |
| Not addicted(ref) | 1 |  |
| **Age** |  |  |
| Adult | 1.50(0.7-3.22) | 0.3 |
| Adolescent(ref) | 1 |  |
| **Sex** |  |  |
| Male | 0.60(0.42-0.87) | 0.008 |
| Female | 1 |  |
| **Faculty** |  |  |
| FESM | 1.56(1.08-2.25) | 0.018 |
| FAAS | 1 |  |
| **Cycle of study** |  |  |
| PhD | 0.89(0.35-2.33) | 0.799 |
| Masters | 0.85(0.58-1.29) | 0.454 |
| Bachelors(ref) | 1 |  |
| **Marital status** |  |  |
| Married | 3.86(0.08-195) | 0.499 |
| Single | 5(0.13-196) | 0.391 |
| Widow | 1.31(0.02-94) | 0.902 |
| Divorced | 1 |  |
| **Religion** |  |  |
| Other | 0.92(0.63-1.32) | 0.642 |
| Catholic | 1 |  |
| **Chronic disease** |  |  |
| Yes | 1.90(1-3.59) | 0.049 |
| No | 1 |  |
| **History of sexual abuse** |  |  |
| Yes | 2.03(0.77-5.37) | 1.52 |
| No | 1 |  |
| **Suicidal ideation** |  |  |
| Yes | 2.69(1.44-5.02) | 0.002 |
| No | 1 |  |

### 3.6. Association between smartphone addiction and anxiety

The odds of anxiety among students with smartphone addiction was 2.44 (1.58-3.77) times the odds of anxiety among students without addiction (p<0.001). After adjusting for age, cycle of study, faculty, marital status, religion, sex, sexual abuse and suicidal ideation, there was still evidence of an association between smartphone addiction and anxiety (Adjusted Odds Ratio: 2.08), p-value: 0.002 (table 5).

**Table 5:** Association between smartphone addiction and anxiety, adjusted for age, sex, faculty, cycle of study, matrimonial status, religion, chronic diseases, sexual abuse and suicidal thoughts.

| Variable | Adjusted Odds Ratio (C.I) | P-Value |
| --- | --- | --- |
| **Addiction** |  |  |
| Addicted | 2.08(1.31-3.30) | 0.002 |
| Not addicted | 1 |  |
| **Age** |  |  |
| Adult | 0.84(0.37-1.92) | 0.684 |
| Adolescent | 1 |  |
| **Sex** |  |  |
| Male | 0.76(0.49-1.16) | 0.204 |
| Female(ref) | 1 |  |
| **Faculty** |  |  |
| FESM | 1.14(0.74-1.73) | 0.558 |
| FAAS | 1 |  |
| **Cycle of study** |  |  |
| PhD | 0.89(0.35-2.23) | 0.799 |
| Masters | 1.07(0.66-1.72) | 0.791 |
| Bachelors | 1 |  |
| **Marital status** |  |  |
| Married | 0.76(0.02-30) | 0.886 |
| Single | 1.39(0.05-38) | 0.845 |
| Widow | 1.26(0.02-66) | 0.909 |
| Divorced | 1 |  |
| **Religion** |  |  |
| Other | 0.95(0.62-1.45) | 0.815 |
| Catholic | 1 |  |
| **Chronic disease** |  |  |
| Yes | 2.32(1.21-4.46) | 0.016 |
| No | 1 |  |
| **History of sexual abuse** |  |  |
| Yes | 1.32(0.49-3.57) | 0.579 |
| No | 1 |  |
| **Suicidal ideation** |  |  |
| Yes | 4.42(2.43-8.00) | <0.001 |
| No | 1 |  |

## 4. Discussion

The objectives of this cross-sectional study were to determine the prevalence of smartphone addiction and common mental disorders and the association between smartphone addiction and these mental disorders among students attending a state university in Cameroon. This study is one of the largest of its kind in sub-Saharan Africa and is an important contribution to the crucial lack of data on mental illnesses and the association between smartphone use and these mental illnesses in sub-Saharan Africa. We found that smartphone addiction, depression and anxiety were prevalent especially among females and bachelor’s degree students. Smartphones were mainly used for social media activities and SMS. There was strong evidence of an association between smartphone addiction and common mental disorders even after controlling for multiple a priori confounders. Apart from the large sample size, other strengths of the study are the use of multistage random sampling to enroll study participants and logistic regression modeling to assess if there is an association between smartphone addiction and common mental illnesses. The above sampling method and the use of logistic regression are key strategies to reduce sampling bias and confounding bias respectively and they both increase the internal validity of the results.

Previous studies have demonstrated that smartphone addiction among students tends to be high [22,23] with some authors reporting addiction percentages of almost 50% in their samples [22]. A previous cross-sectional study involving medical students in multiple state universities in Cameroon reported major depressive disorders in about a third of study participants [24]. Our results are comparable to these findings and our study further concurs with this previous report in that students with symptoms of common mental disorders tended to be females. Larger and more robust studies in the United Kingdom [25] and the United States of America [26] concluded that women have higher rates of common mental disorders than men. It has been proposed that the higher prevalence of depressive symptoms in women may be as a result of triggering by hormonal changes, particularly during puberty, prior to menstruation, following pregnancy and at perimenopause [27]. Mental health improvement programmes must consider this gender difference in order to foster equity in the access to and utilisation of effective mental health services.

Social media and smartphones took most of the time spent using smartphones. Similar results were obtained in previous studies conducted out of Africa [28,29] which found that social networking are the most time spending activities. Thus, our results concur with the general tendency for problematic smartphone users to report social media as the most preferred activity on smartphone [27]. In the general population, smartphone and social media use are far more common in younger and educated population segments as our study sample.

The magnitude of the association (when observed) between problematic smartphone use and common mental disorders varies across literature and this may be due to the differences in data collection methods (such as face-to-face versus online), and assessment tools for measuring addiction and mental health disorders, and variations in baseline characteristics of the study populations. For instance, our study involved a student population that was relatively young, and the data was collected via face-to-face interviews while in the Middle East, Alhassan and colleagues studied data collected using web-based questionnaires administered to adults [29,30]. The two studies also used different methods of measuring depressive symptoms: Beck’s Depression Inventory in the study by Alhassan and collaborators [17] and PHQ-9 in our study. A global review by Sohn and colleagues found that there is a significant association between problematic smartphone use and common mental health disorders [17]. Albeit our results concur with the findings of this review, it should be noted that the review synthesised evidence from Europe, America and Asia and not Africa [17]. Further studies in the sub-Saharan African context are therefore needed to make more meaningful comparisons.

This study is not without limitations. Data were collected from students from the university of Dschang, so the results should be interpreted with caution when referring to other segments of the general population. Additionally, social desirability bias and recall bias during the filling of the questionnaire as well as lack of adjustment for other potential confounders such as socioeconomic status, sleep pattern and history of intimate partner violence may have reduced the validity of our findings. Moreover, it is possible that the validity of the results was also affected by measurement bias given that depression and anxiety were assessed using self-reported psychological symptoms measured using PHQ-9 and PHQ-7 which have not been validated in Africa. In order to provide much stronger evidence on the association between smartphone use and common mental illnesses, larger observational studies involving structured interviews by clinicians, adjustment for a broader range of confounders and assessment of reverse causality should be conducted.

## 5. Conclusion and recommendation

Depression and anxiety were prevalent in the study population. Smartphone addiction was found to be associated with these common mental disorders. In this setting, it is crucial to foster strategies of prevention and promotion in mental health especially among problematic smartphone users. Apart from raising awareness on these issues among students, the ministry of higher education in collaboration with the ministry of health could consider integrating more functional psychosocial support and basic mental health services with pre-existing programmes in university settings. In order to assess if there is evidence of stronger causal relationships between smartphone addiction and mental health problems, much larger observational studies are warranted.

## Data Availability

The data used to support the findings of this study are available from the corresponding author upon request.

## Abbreviations

CI: Confidence Interval
FAAS: Faculty of agronomy and agricultural sciences
FESM: Faculty of economic sciences and management
FLHS: Faculty of Letters and Human Sciences
FLPS: Faculty of Law and Political Sciences
FS: Faculty of Science
LMICs: Low and middle-income countries
PHQ-7: Patient health questionnaire 7
PHQ-9: Patient health questionnaire 9
SAS-SV: Smartphone addiction scale short version
SMS: Short Message Service

## Authors’ contributions

Martial Pianta Sonkoue and Jerome Ateudjieu conceived the study and did the literature review. They also prepared the protocol for this study. Martial Pianta Sonkoue, Miranda Baame Esong and Cyrielle Djouda Douanla collected the data. Martial Pianta Sonkoue, Benjamin Momo Kadia and Jerome Ateudjieu analysed and interpreted the data. Martial Pianta Sonkoue, Miranda Baame Esong and Cyrielle Djouda Douanla prepared and edited the manuscript. Benjamin Momo Kadia critically reviewed the technical and intellectual consistency of the manuscript. All authors read and approved the final manuscript.

